# Predicting normative walking biomechanics across the lifespan using seven simple features

**DOI:** 10.1101/2025.03.19.25324241

**Authors:** Bernard X.W. Liew, Rachel Senden, David Rugamer, Emanuel Sommer, Kenneth Meijer, Qichang Mei, Richard Foster, Matthew Taylor

## Abstract

**Objectives:** The assessment of gait impairments requires a normative reference for comparison. For a fair assessment, comparisons must be made against a reference standing after controlling for sex, anthropometry, and walking characteristics. This study aimed to develop statistical models that predict the lower-limb kinematics and kinetics of walking across the lifespan of healthy participants, using seven simple covariates. Sixteen statistical models predicted 16 joint kinematics and kinetics during walking using the covariates of sex, age, height, mass, side (laterality), walking speed, and cadence, which were developed based on 301 participants between three to 91 years old. The root mean squared error (RMSE) ranged from 4.71° to 7.97° for joint angles, within 0.07N/kg for ground reaction forces, 0.09 to 0.15 Nm/kg for joint moments, and 0.33 to 0.39 W/kg for joint powers. We provide both online and local apps which can be easily used by clinicians and scientists to generate normative walking data with uncertainty values, which can be used for movement impairment analysis (https://github.com/EmanuelSommer/ShinyFOSR).

## Introduction

Movement impairments are common in many neurological [1], musculoskeletal [2], cardiovascular [3] disorders, and even manifest naturally due to ageing [4]. The clinical assessment of movement impairments in patients requires the comparison with typical reference standards from a healthy person or cohort [5, 6]. To ensure a correct interpretation, comparisons must be made with healthy controls after controlling for sex, anthropometry (e.g. body mass index, height), and spatiotemporal walking characteristics (e.g. walking speed) [7].

The most common method to compare movement impairments is based on tools, termed “gait indices”, such as the Gait Deviation Index [8–10], Gait Profile Score [11], and the Gillette Gait Index [8]. A challenge when using traditional gait indices is that it requires clinicians or researchers to collect data on individuals defined as “normative”, from scratch to develop a database upon which a normative index is quantified and compared. This process can be very time-consuming and expensive. To our knowledge, no open-source “normative” dataset or normative gait indices has been published to enable researchers to compare. Another limitation of traditional gait indices is that typical movements can be affected by movement-specific factors, like the speed of walking [12], and potentially individual-specific factors, like age. This means that if the gait index was developed based on a sample with a different characteristic from the target patient, or performed a movement with different task requirements, traditional gait indices may classify the target patient as impaired.

An alternative to the use of traditional gait indices is a direct population-based statistical approach towards quantifying an average with a measure of uncertainty of any biomechanical parameters of interest [7, 13, 14]. Population-based statistical approaches in biomechanics, such as Statistical Shape Modelling, are not new and have been largely used to scale the morphology of differential anatomical region(s) from a generic model to fit the anthropometry of the measured individual [15]. Fewer research has focused on the statistical prediction of normal human motion, particularly via predictive factors that are simple, quick, and cheap to measure clinically. A previous study reported that the sagittal plane joint angles during walking of the ankle, knee, and hip could be predicted within <2°, based only on knowing an individual’s walking speed, gender, age, and body mass index [7]. Another study reported that lower-limb joint angles can be predicted within a root-mean-squared error (RMSE) of 4.45° to 6.61° based on age, height, and weight, or based on the shape of the lower-limb bones [16].

A limitation of prior studies [7, 16] is the inclusion of a restricted age spectrum of participants (e.g. 39 ± 13 years in [7], and 22 ± 2 years in [16]), the assumption of statistical linearity between the covariates (also known as predictors) and the biomechanical outcomes and the inclusion only of kinematic features as outcomes for prediction. It is well known that kinematic and kinetic gait differences can be observed across the human lifespan [17]. For example, in a study, only children reported that ankle push-off power (A2) appeared to mature by 4 years old [18]. Separate research only on adults (i.e. ≥ 18 years old) reported that A2 power peaked again at 60 years [17]. A population-based statistical model of gait biomechanics must include participants from a wider age spectrum of young children to very old adults (i.e. 80+ years). Previous studies also assumed a linear relationship between the included covariates and biomechanical outcomes using statistical techniques like partial least squares regression [16] and linear stepwise regression [7]. However, previous studies have reported nonlinear relationships between covariates such as age and biomechanical features like joint power [17]. In other words, the association between a covariate and a predictor varies depending on the value of the covariates.

This study aims to develop statistical models that predict the lower-limb kinematics and kinetics of walking across the lifespan of healthy participants, using simple covariates that can be easily measured in clinical settings, such as sex, age, height, mass, side (laterality), walking speed, and cadence. We hypothesised that the root-mean-squared error (RMSE) for lower-limb joint angles to be <7° [16], based only on the covariates of sex, age, height, weight, side, and cadence. For joint kinetics, given that no prior studies have reported RMSE values based on prediction models using sex, age, height, weight, side, and cadence as covariates, we hypothesised that our predicted joint moments will have an RMSE of < 0.4 Nm/Kg [19].

## Methods

### Participants

Normative data collected from previous studies of Senden et al were used for this study, and the two studies included 55 typically developed children [20] and 246 healthy adults [21].

### Data collection

Three-dimensional (3D) gait analysis was performed at the motion lab of the MUMC+ using the Computer Assisted Rehabilitation Environment (CAREN, Motek Medical BV, Amsterdam) system. Participants walked on an instrumented split-belt treadmill (ForceLink, Culemborg, 1000Hz), while marker trajectories were captured with a 12-camera optical motion capture system (Vicon, Oxford, 100 Hz). The Human Body Lower Limb Model (HBM-II) was used as a biomechanical model [22]. Participants wore standardised gymnastic shoes and a safety harness to prevent falls.

After a six-minute familiarisation period at comfortable walking speed, participants walked for 250 steps at a comfortable, slow (30% slower than comfortable), and fast (30% faster than comfortable) speed, which were performed in a random order. To determine comfortable walking speed, children performed repeated overground walking trials over a nine-meter walkway, while speed was measured using two movement detection ports. For the healthy adults, the RAMP protocol was used where subjects started to walk on the treadmill at 0.5m/s while the speed was gradually increased to 0.01m/s every second until a comfortable speed was reached. This was repeated three times and the average of three repetitions was used as the comfortable speed.

The force plate configurations for the analog data were set at 10 Hz for the low-pass prefilter frequency and 20 N for the force threshold. Marker trajectories and force plate data were filtered with a unidirectional 2^nd^ order Butterworth filter at 6 Hz. Gait event detection was defined based on a combination of heel marker kinematics and force plate data (exceeding the threshold of 50 N) [23]. Custom Matlab scripts were used to check data quality and calculate spatiotemporal parameters, kinematics, and kinetics. Kinematic and kinetic data were time-normalised to 100 data points between two consecutive initial contacts for each side. GRF, joint moment and power data were subsequently normalised to body mass (kg). The kinematic and kinetic trajectories have been uploaded and are freely available in the Open Science Framework (OSF) (children: https://osf.io/3xqew/; adults: https://osf.io/t72cw/).

### Statistical analysis

All analyses were conducted in R software (v4.4.2) using the *refund* package (v0.1-37). We developed 16 statistical models to predict 16 kinematic and kinetic trajectories – angles (hip flexion, abduction, rotation; knee flexion; ankle plantarflexion and inversion), GRF (vertical and anterior-posterior), moments (hip flexion, abduction, rotation; knee flexion; ankle plantarflexion), and powers (hip, knee, ankle flexion). For all models, the same covariates were included: sex (male or female), age (years), walking speed (m/s), body mass (kg), height (m), cadence (steps/min), side (left or right), and the random effect of the subject. We use a function-on-scalar model for each of the three joints – specifically the *pffr* function in the *refund* package. As a functional additive model, we model the expectation of each outcome trajectory yi(t) for every subject i at gait point t.

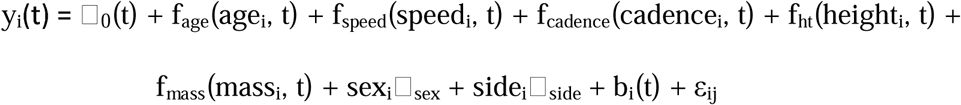

Where □_0_(*t*) is the model’s time-varying intercept,□_sex_ a time-independent effect of sex, □_side_ a time-independent effect of side, f( , *t*) indicate functional effects that are estimated to be non-linear both in the direction of the covariate and across time, bi(*t*) are time-varying random intercepts, and ε_i_ and independent Gaussian error term.

An 80:20 split of the data was performed to create a training set (80%, n = 240 participants) to develop the models, and a testing set (n = 61 participants) to validate the accuracy of the outcome predictions. The prediction performance of the models was determined by comparing the 16 predicted outcomes in the test set, against their original values using the Root Integrated Mean Squared Error (BW), relative Root Integrated Mean Squared Error (relRMSE, %) [24], and Pearson correlation coefficient (cor) [25, 26].

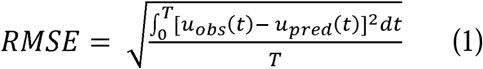

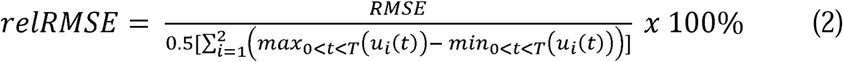

where *T* represents the stance duration between initial contact and toe-off, *u_obs_*(*t*) represents the value at the *t^th^* time point of the observed outcome, *u_pred_*(*t*) represents the value at the *t^th^* time point of the predicted outcome, and *i* represents either the observed or predicted outcomes. We also report the association (point estimate and 95% confidence interval [CI]) between the outcomes and each of the fixed effect covariates. For the continuous covariates of age, speed, cadence, height and mass, we selected the 25^th^, 50^th^, and 75^th^ quantiles of the values to plot the smooth association estimate.

To make the model open source and easily accessible, we retrained the models using the entire dataset (n = 301) and created a shiny app (https://github.com/EmanuelSommer/ShinyFOSR). for readers to input different covariate values to predict the mean waveform value of different kinematic and kinetic outcomes. Readers can use the app to predict, plot, and export the values as a figure and table. We also included a reference with a detailed and easy set of instructions to download and use the app locally on their computer. This local version can predict the mean values and the 95%CI or the standard deviation.

## Results

Basic descriptive summaries of the cohort can be found in Figure 1. Figures 2 and 3 illustrate the average predicted kinematic and kinetic trajectories, alongside the observed trajectories in the test dataset. RMSE ranged from 4.71° to 7.97° for kinematic data, while relRMSE ranged from 9.85% to 50.72% (Table 1). Accuracy was the best generally for sagittal plane joint angles and the poorest for transverse plane angles (Table 1). For kinetic data, the statistical models predicted GRFs to be within 0.07N/kg and <7% for RMSE, and relRMSE (Table 1). The statistical models were better at predicting joint moments than joint powers, with an accuracy for the former ranging from 0.09 to 0.15 Nm/kg for RMSE and 9.23% to 24.71% for relRMSE; and for the latter ranging from 0.33 to 0.39 W/kg for RMSE and 12.98 to 17.91% for relRMSE (Table 1).

**Figure 1.**
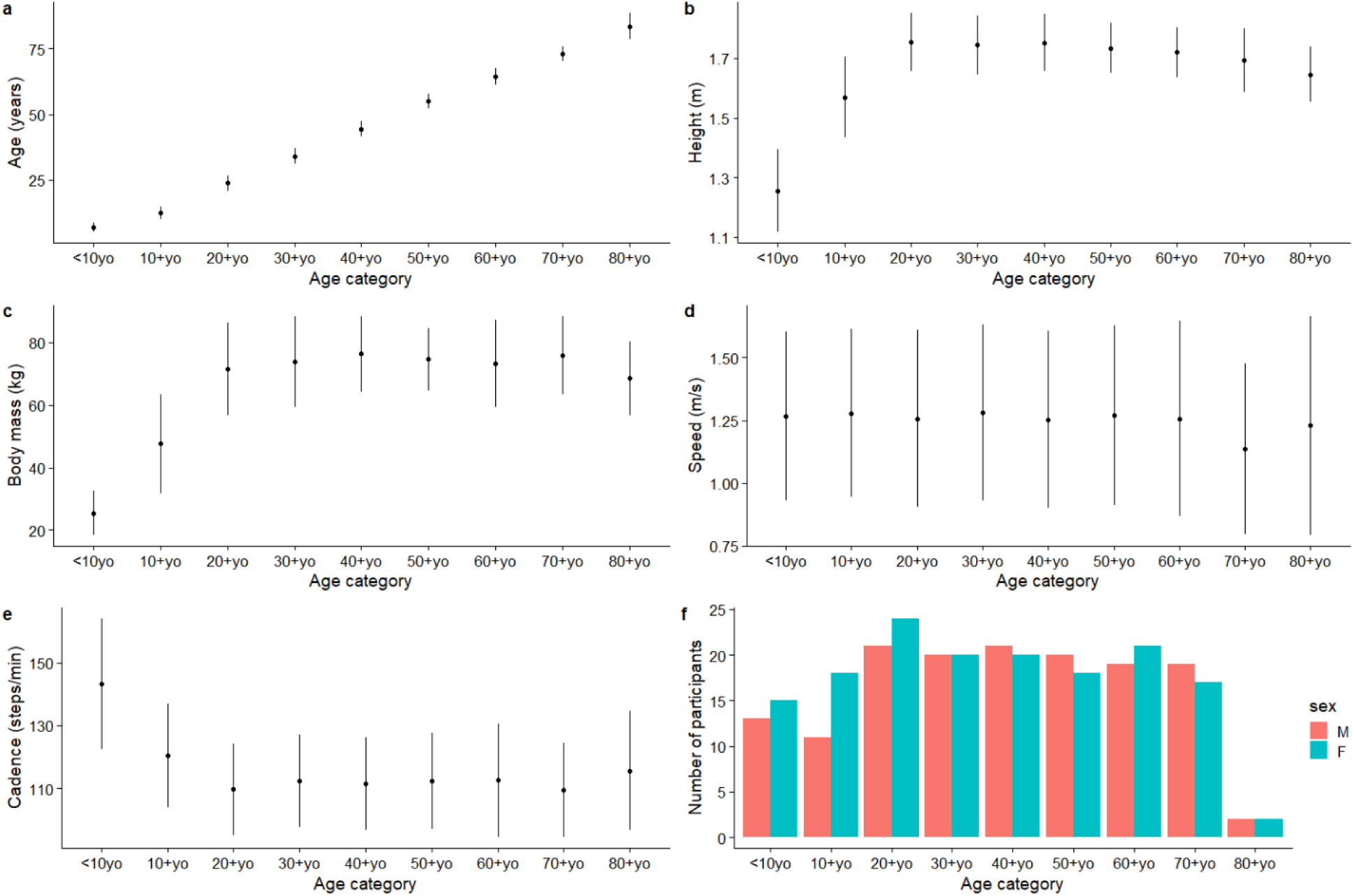
Descriptive characteristics of the included participants (n = 301). Point estimate reflect means with error bars representing one standard deviation (a – e), apart from the number of participants (f).

**Figure 2.**
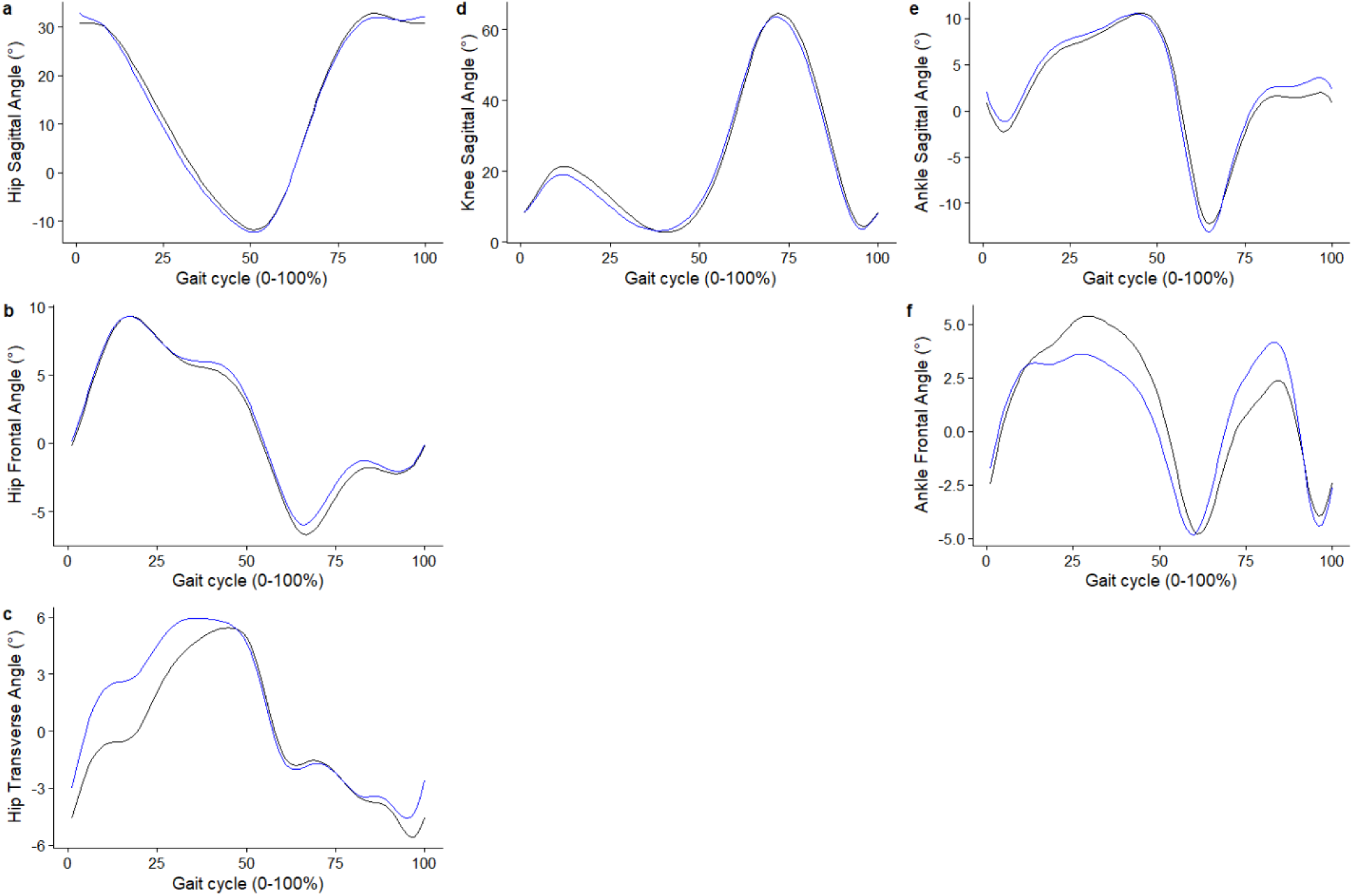
Average predicted (blue) kinematic trajectories against the observed (black) trajectories for the test dataset.

**Figure 3.**
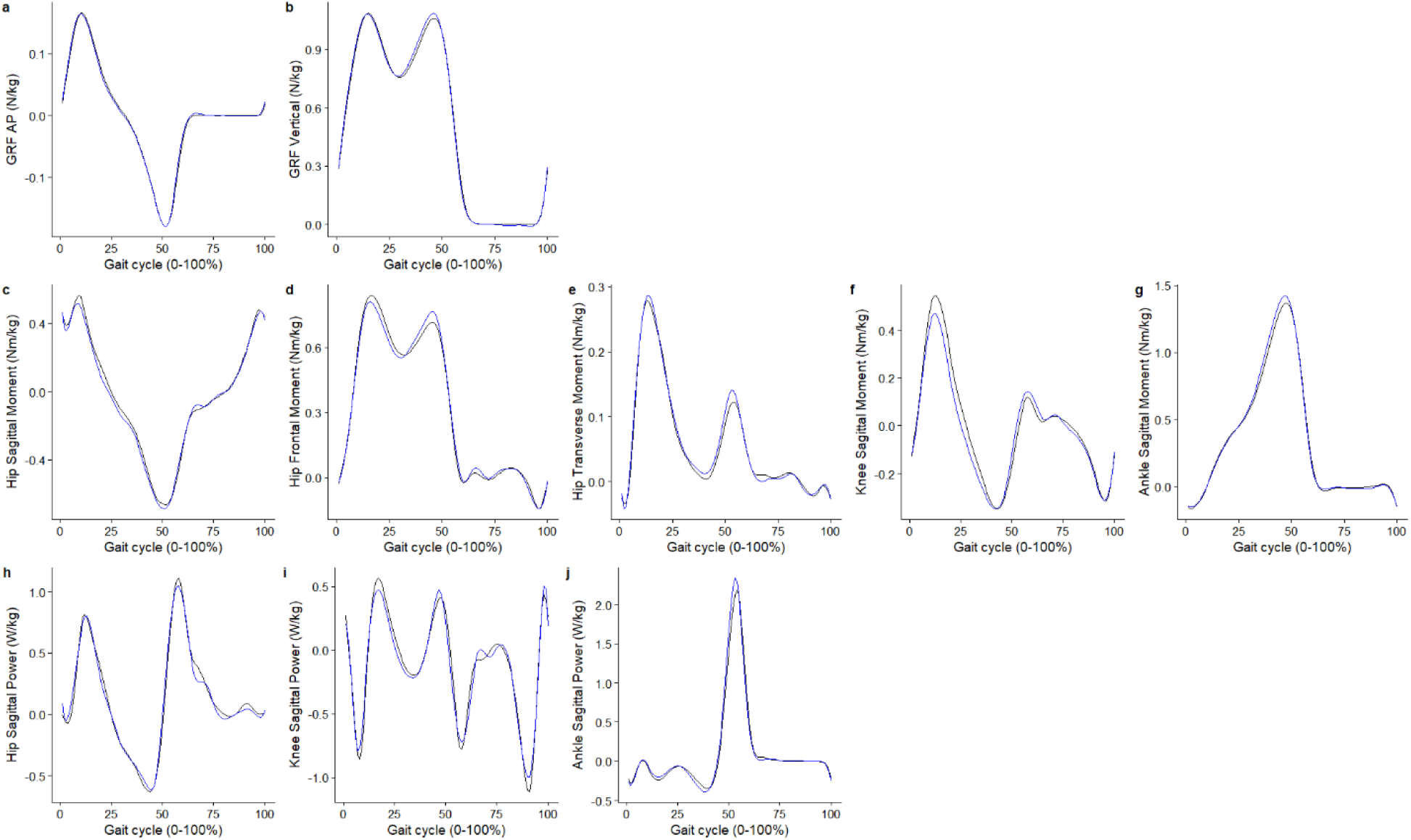
Average predicted (blue) kinetic trajectories against the observed (black) trajectories for the test dataset.

**Table 1.**
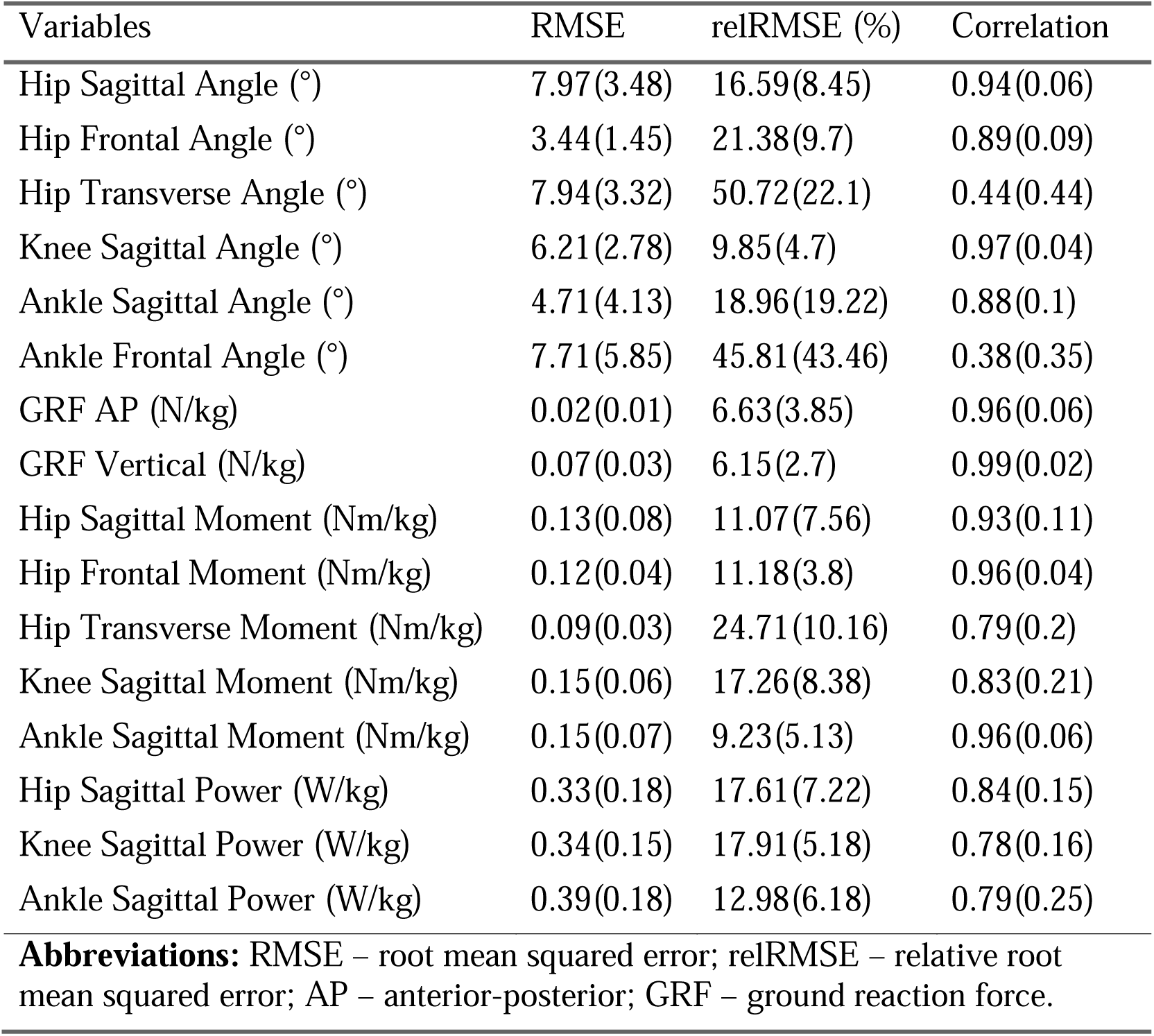
Accuracy of predicted kinematic and kinetic trajectories.

| Variables | RMSE | relRMSE (%) | Correlation |
| --- | --- | --- | --- |
| Hip Sagittal Angle (°) | 7.97(3.48) | 16.59(8.45) | 0.94(0.06) |
| Hip Frontal Angle (°) | 3.44(1.45) | 21.38(9.7) | 0.89(0.09) |
| Hip Transverse Angle (°) | 7.94(3.32) | 50.72(22.1) | 0.44(0.44) |
| Knee Sagittal Angle (°) | 6.21(2.78) | 9.85(4.7) | 0.97(0.04) |
| Ankle Sagittal Angle (°) | 4.71(4.13) | 18.96(19.22) | 0.88(0.1) |
| Ankle Frontal Angle (°) | 7.71(5.85) | 45.81(43.46) | 0.38(0.35) |
| GRF AP (N/kg) | 0.02(0.01) | 6.63(3.85) | 0.96(0.06) |
| GRF Vertical (N/kg) | 0.07(0.03) | 6.15(2.7) | 0.99(0.02) |
| Hip Sagittal Moment (Nm/kg) | 0.13(0.08) | 11.07(7.56) | 0.93(0.11) |
| Hip Frontal Moment (Nm/kg) | 0.12(0.04) | 11.18(3.8) | 0.96(0.04) |
| Hip Transverse Moment (Nm/kg) | 0.09(0.03) | 24.71(10.16) | 0.79(0.2) |
| Knee Sagittal Moment (Nm/kg) | 0.15(0.06) | 17.26(8.38) | 0.83(0.21) |
| Ankle Sagittal Moment (Nm/kg) | 0.15(0.07) | 9.23(5.13) | 0.96(0.06) |
| Hip Sagittal Power (W/kg) | 0.33(0.18) | 17.61(7.22) | 0.84(0.15) |
| Knee Sagittal Power (W/kg) | 0.34(0.15) | 17.91(5.18) | 0.78(0.16) |
| Ankle Sagittal Power (W/kg) | 0.39(0.18) | 12.98(6.18) | 0.79(0.25) |
| **Abbreviations:** RMSE – root mean squared error; relRMSE – relative root mean squared error; AP – anterior-posterior; GRF – ground reaction force. |  |  |  |

## Smooth effect plots

The smooth effect plots support the nonlinear association between the included covariates of age and speed and the kinematic and kinetic outcomes. For the covariate age, the greatest effect is on the hip sagittal plane angle at 26% of the gait cycle, where at the age of 46 years it was associated with greater hip extension angle by -16.9° (95%CI -17.3° to -16.5°) (Figure 4). For joint moments, age had the greatest effect on the hip sagittal moment, where at age 46 years, it was associated with greater hip flexion moment by -0.35Nm/kg (95%CI -0.36 to -0.33Nm/kg) at 21% of the gait cycle (Figure 5). Age also had the greatest effect on hip flexion power, where at age 46 years, it was associated with greater hip power absorption by -0.35W/kg (95%CI -0.39 to -0.31W/kg) at 19% of the gait cycle (Figure 5).

**Figure 4.**
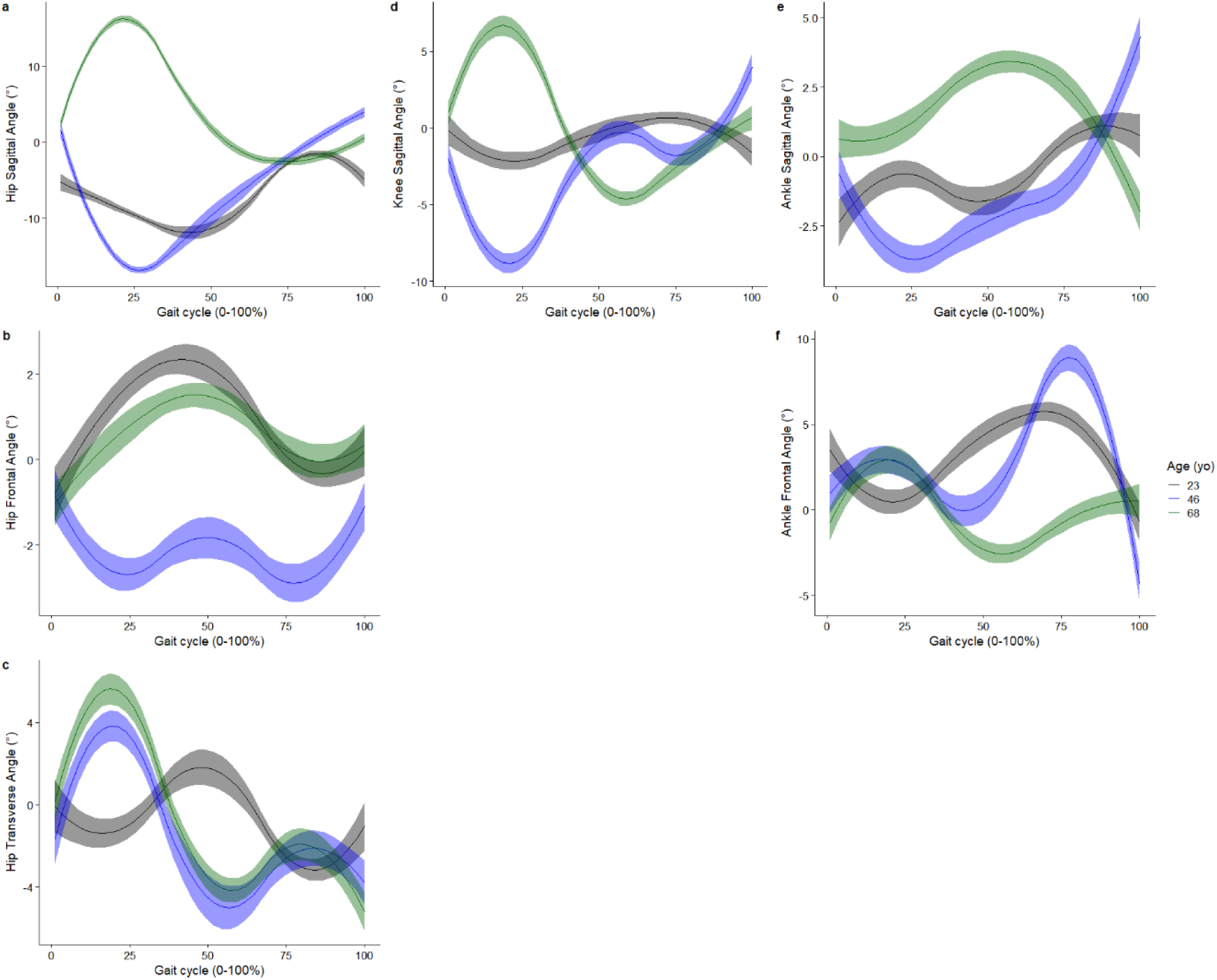
Smooth plots with error clouds as 95% confidence interval for the covariate of age and joint kinematics.

**Figure 5.**
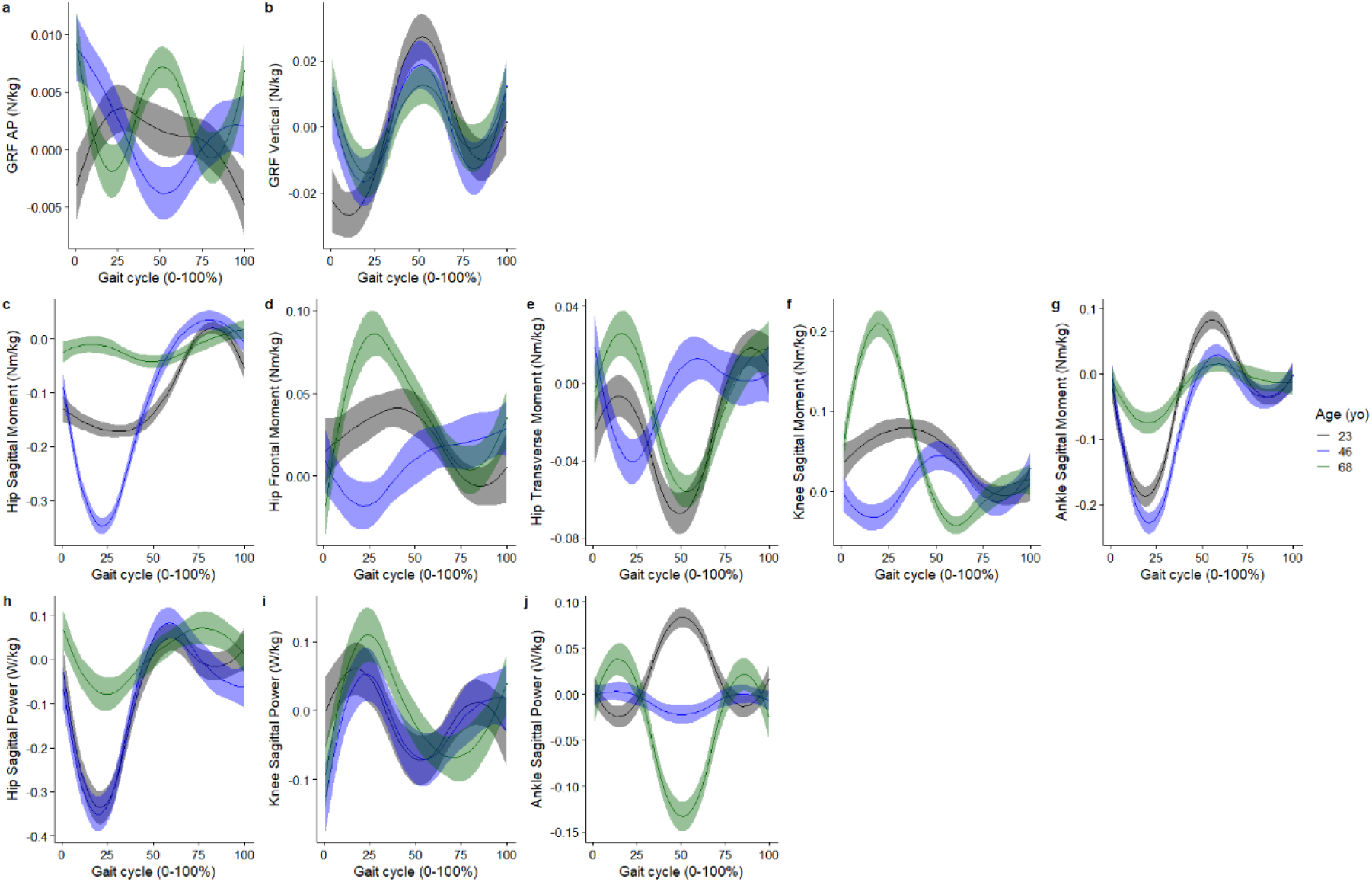
Smooth plots with error clouds as 95% confidence interval for the covariate of age and joint kinetics.

For the covariate speed, the greatest effect is on the hip sagittal angle at 1% of the gait cycle, where with a speed of 1.84m/s, it was associated with an increase in hip flexion angle 13.4° (95%CI 13.4° to 14.2°) (Figure 6). For joint moments, speed had the greatest effect on the hip sagittal moment, where at a speed of 1.84m/s, it was associated with an increase in hip extensor moment by 0.43Nm/kg (95%CI 0.42 to 0.45Nm/kg) at 1% of the gait cycle (Figure 7). Speed had the greatest effect on ankle plantarflexion power, where at a speed of 1.84m/s it was associated with an increase in power generation by 0.82W/kg (95%CI 0.79 to 0.85W/kg) at 49% of the gait cycle (Figure 7).

**Figure 6.**
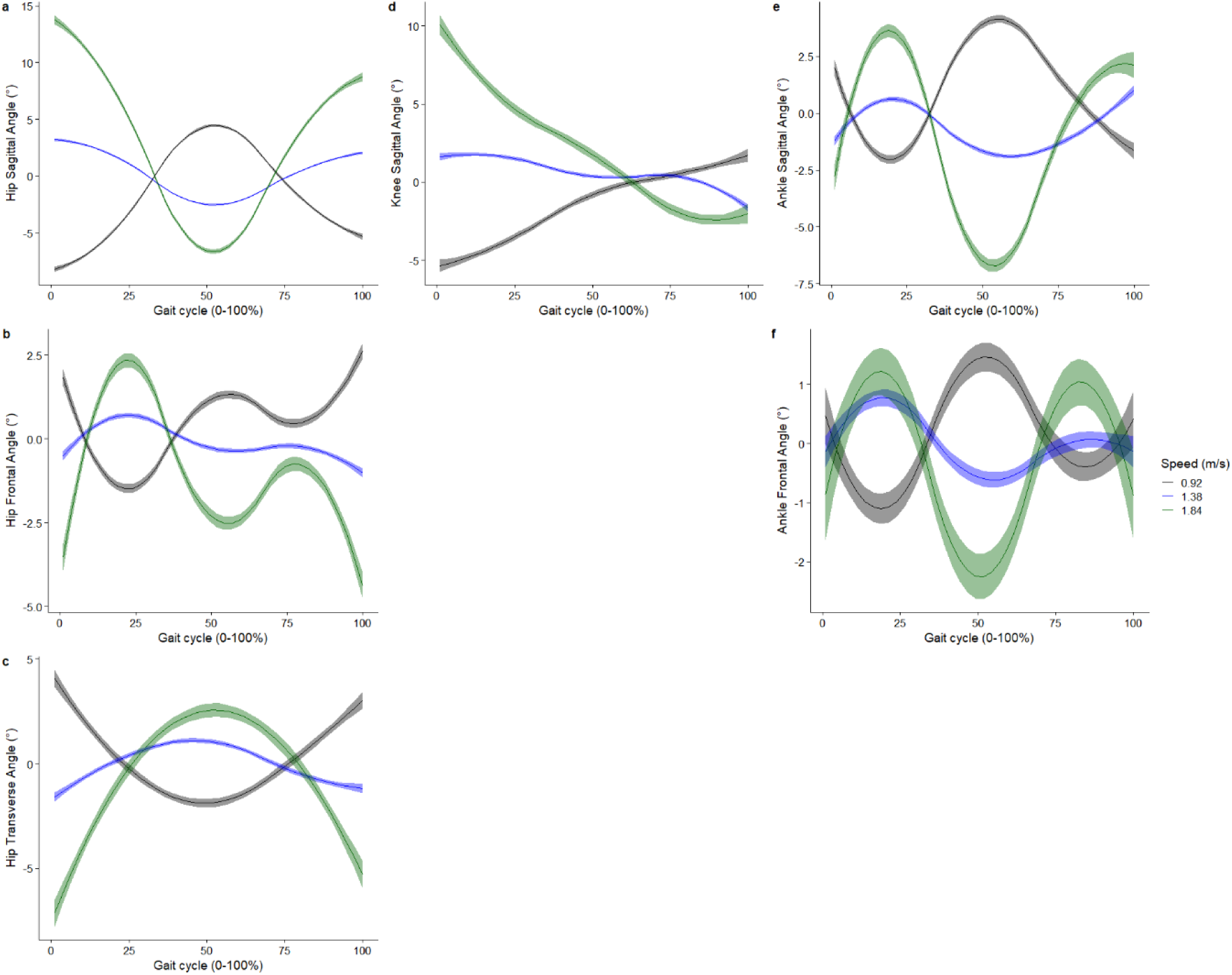
Smooth plots with error clouds as 95% confidence interval for the covariate of speed and joint kinematics.

**Figure 7.**
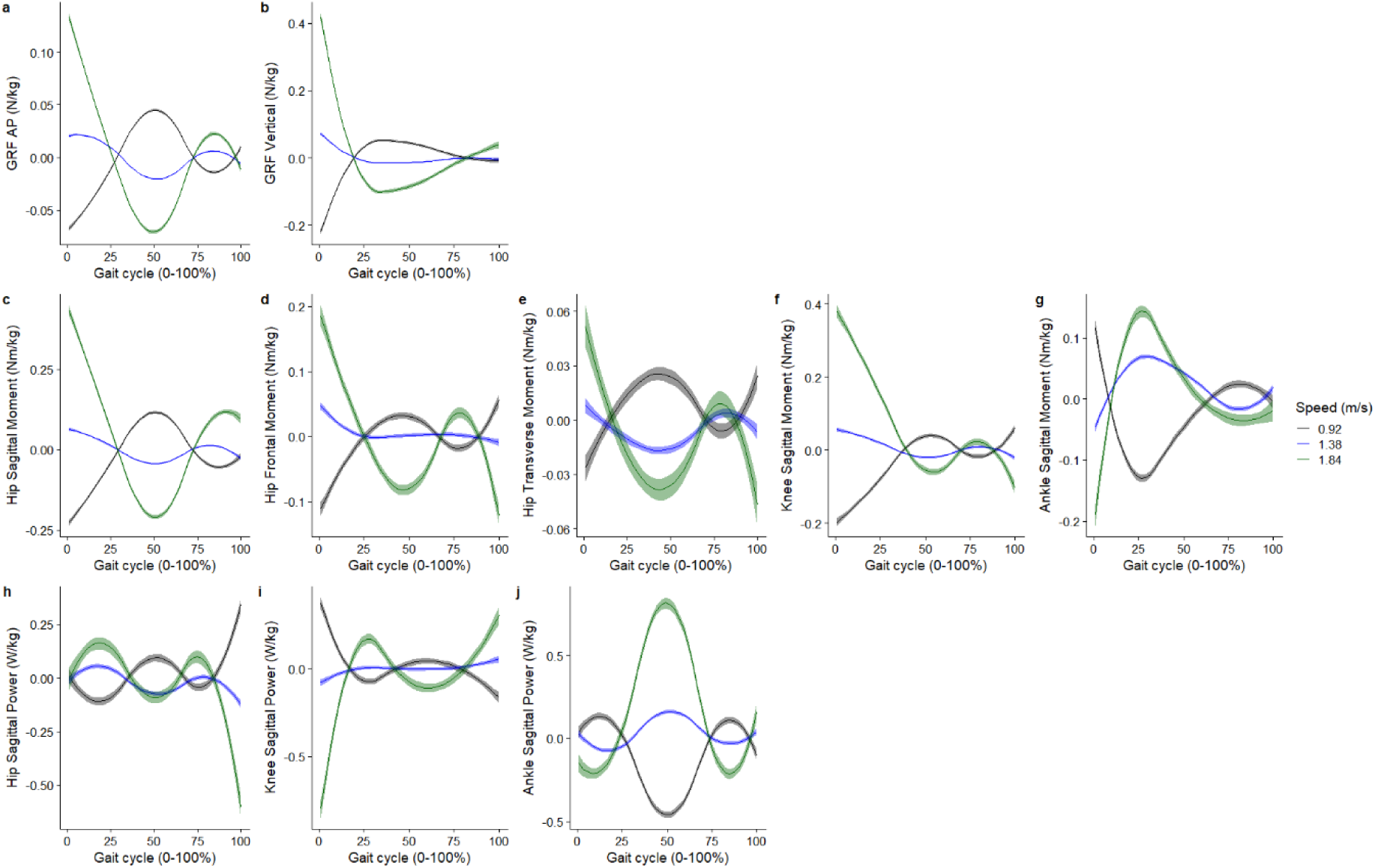
Smooth plots with error clouds as 95% confidence interval for the covariate of speed and joint kinetics.

## Discussion

Determination of impaired movement depends on knowing normal movement. Statistical modelling based on simple clinical measures represents a simple way that scientists and clinicians can predict key walking biomechanics of healthy individuals. The RMSE averaged across the stride cycle for the lower-limb joint angles was <8°, only marginally poorer than our hypothesis of <7° [13]. The RMSE was between 0.09 to 0.15 Nm/kg for joint moments, which was better than previously predicted joint moments of < 0.4 Nm/kg using joint angles as predictors [16].

The predicted RMSE for joint angles in this study was approximately 5° greater than previous studies which reported that RMSE for sagittal plane joint angles was within 2° [7, 27]. Previous studies have reported that the minimal detectable change values of lower-limb joint angles during walking of up to 7°[28]. RMSE values greater than minimal detectable change values suggest the accuracy of some of the predictive models presently may be insufficient to be used clinically. However, even though the error metrics presented presently reflect the waveform-averaged, the accuracy of our prediction models is not uniform across the stride cycle as observed in Figures 2 and 3, with better accuracy in some phases of the stride cycle than in other phases. Users of the present prediction models in defining normative walking gait biomechanics should be aware of our reported RMSE values, and determine clinically if these errors exceed a clinically meaningful threshold.

The differences in the predicted accuracy of our biomechanical waveforms between studies may be due to different validation methods [7, 27]. Previous studies used leave-one-subject-out cross-validation (LOSOCV) as their validation approach [7, 27], whereas the present study used a train-test split approach. LOSOCV uses more data, apart from one subject, for training. This means that a greater percentage of the data was used for training, and a smaller percentage of data was used to validate the model, compared to a train-test split validation approach. For example, the study of Moissenet et al. [7] used 1325 observations (53 subjects, 5 speeds, 5 trials) for each training loop, whereas we used only 720 observations (240 subjects, 3 speeds) for training. The present findings thus provide a more conservative estimate of gait kinematics predictive modelling, that may be more transferrable to unseen real-world applications.

The present study demonstrates that normative joint kinetics during walking can be predicted using simple clinical measures, without having to rely on more complex biomechanical inputs as predictors. The predicted RMSE for GRF, moments, and powers in this study ranged between 0.02 to 0.07N/kg, 0.09 to 0.15Nm/kg, and 0.33 to 0.39 W/kg, respectively. Surprisingly, these error ranges were less than machine learning prediction models of joint moments using 3D motion capture inputs in running (RMSE: 0.07 to 0.54Nm/kg)[19], and less than models using inertial measurement units (IMUs) as inputs for walking (RMSE: 0.07 to 0.24 Nm/kg) [29]. Wouda et al. predicted vertical GRF to a RMSE of < 0.27BW using three IMU inputs for running [30]. Another study predicted ankle joint power for walking using IMU sensors with a RMSE of <0.21 W/kg [31]. Statistical models like the present and others [7], are useful for population average prediction – which cannot capture variation between movement cycles, whereas more complex prediction models built upon biomechanical input predictors may be more useful for capturing cycle-specific predictions.

Whilst the present study did not specify apriori a linear relation, previous studies have assumed a linear relationship between the predictors investigated (e.g. age, speed) and the biomechanical outcomes [7, 14], whilst another study assumed a quadratic association with the outcomes [27]. The present study reported a non-linear association between age and ankle plantar flexion, given that ankle plantar flexion increased between 23 to 48 years old and decreased from 48 to 68 years old (Figure 4e). The non-linear association between age and our 16 biomechanical outcomes investigated was evident in our smooth plots (Figures 4, 5). Studies which used linear regression methods reported non-significant linear associations between age and ankle plantar flexion during push-off [7], while other studies reported a decreased ankle plantar flexion during push-off in older adults compared to younger adults [32]. This study provides evidence that it may not be appropriate to assume linearity across all values of the covariates and the outcome across a gait cycle. Hence, whilst linear-regression-based approaches provide very interpretable results (e.g. a beta coefficient), they may not adequately model the relationships between the covariates and the biomechanical outcomes.

A strength of the present study is the fact that we included 301 participants between three and 91 years old, which represents a five times greater sample size than recent studies [7]. Apart from the age category of >80 years old, the current study included >20 participants in each decade of years for adults, included in the modelling, making our statistical model more representative of a lifespan population cohort. Another advantage of this study was the inclusion of simple clinically oriented variables as covariates, which will enable immediate clinical translation to derive a normative understanding of walking biomechanics. However, this study has also some limitations. First, the included participants were collected whilst walking on a treadmill, which might differ from overground walking [33]. Differences between treadmill and overground walking ranged from 0.84° (pelvic tilt) to 6.42° (peak knee flexion during swing) for joint angles, and 0.02 Nm/kg (peak knee extension moment) to 0.32 Nm/kg (peak hip extension moment) for joint moment[33]. Second, although we included covariates which previously have been shown to have a significant association with walking biomechanics, more variables may play an important role when developing population-based models which were currently not measured, like ethnicity [34] and psychological factors [35]. Future studies may explore this further.

The present predictive models could be used like other more established tools like the Gait Deviation Index [8–10], to provide a benchmark against which, impaired movements can be compared. Unfortunately, tools like the Gait Deviation Index require researchers to collect data on healthy individuals from scratch to quantify the index. Also, how the Gait Deviation Index is quantified means that it is not specific to key personal and gait characteristics. In contrast, the availability of our prediction models via online or our local app enables end-users to quickly generate both the mean and uncertainty values, against which impaired movements can be compared.

## Conclusions

Statistical models using seven clinical covariates can predict most normative walking kinematics and kinetics, apart from non-sagittal plane kinematics, to a level comparable to more complex machine learning models using biomechanical covariates. Both our online and local apps can be used by clinicians and scientists to generate normative data with uncertainty values, which can be plotted against patient data for reporting.

## Data Availability

All data produced in the present study are available upon reasonable request to the authors.

